# A Comparison of Aerosol Mitigation Strategies and Aerosol Persistence in Dental Environments

**DOI:** 10.1101/2021.07.30.21261399

**Authors:** Shruti Choudhary, Michael J. Durkin, Daniel C. Stoeckel, Heidi M. Steinkamp, Martin H. Thornhill, Peter B. Lockhart, Hilary M. Babcock, Jennie H. Kwon, Stephen Y. Liang, Pratim Biswas

**Author notes:** Primary corresponding author. Alternate corresponding author.

## Abstract

**Objectives:** To determine the impact of various aerosol mitigation interventions and establish duration of aerosol persistence in a variety of dental clinic configurations.

**Methods:** We performed aerosol measurement studies in endodontic, orthodontic, periodontic, pediatric, and general dentistry clinics. We used an optical aerosol spectrometer and wearable particulate matter sensors to measure real-time aerosol concentration from the vantage point of the dentist during routine care in a variety of clinic configurations (e.g, open bay, single room, partitioned operatories). We compared the impact of aerosol mitigation strategies [ventilation and high-volume evacuation (HVE)] and prevalence of particulate matter in the dental clinic environment before, during and after high-speed drilling, slow speed drilling and ultrasonic scaling procedures.

**Results:** Conical and ISOVAC® HVE were superior to standard tip evacuation for aerosol-generating procedures. When aerosols were detected in the environment, they were rapidly dispersed within minutes of completing the aerosol-generating procedure. Few aerosols were detected in dental clinics – regardless of configuration – when conical and ISOVAC® HVE were used.

**Conclusions:** Dentists should consider using conical or ISOVAC® HVE rather than standard tip evacuators to reduce aerosols generated during routine clinical practice. Furthermore, when such effective aerosol mitigation strategies are employed, dentists need not leave dental chairs fallow between patients as aerosols are rapidly dispersed.

**Clinical Significance:** ISOVAC® HVE is highly effective in reducing aerosol emissions, with adequate ventilation and HVE use, dental fallow time can be reduced to 5 minutes.

## 1. Introduction

Certain dental procedures generate high concentrations of aerosols in a patient’s mouth, which may increase the risk for transmission of respiratory pathogens to dental healthcare professionals (DHP) and other patients. Most aerosols are generated and emitted from two sources: (a) high-speed, high-frequency dental instruments that use water for cooling, and (b) the patient’s bodily fluids, including blood, saliva, dental plaque, and food debris [1].

Previous studies show that dentists and patients are exposed to greater than 10,000 bacteria/m^3^, and the risk of inhaling infectious aerosols during dental procedures may be high [2,3]. It is estimated that dentists and other DHPs may inhale up to 0.014 µl of saliva over a 15-minute peak exposure period, and up to as much as 0.12 µl of saliva in extreme instances [4]. Several studies have examined emissions during dental procedures. Ultrasonic scaling has been associated with an increased burden of bacterial contamination [5,6]. In dental clinics, aerosols can remain prevalent in the surrounding environment for as long as 30 minutes after a procedure has been performed [7,8,9]. However, these studies only provide daily average values, did not take into account the impact of aerosol mitigation interventions, and did not measure exposure from the vantage point of the dentist or other DHPs [10,11].

Preliminary data derived from mannequin experiments suggests that the risk posed by aerosols can be reduced with high-volume evacuation (HVE), rubber dam isolation, flushing water lines at the beginning of the work day and between patients, pre-procedural patient mouth rinses, and appropriate use of personal protective equipment by DHP [1,12,13,14,15]. However, these studies were not performed during actual patient care and lack real-time particle monitoring necessary to determine the effectiveness of these interventions.

The objectives of this study were to use real-time optical aerosol instruments to 1) characterize aerosol emissions from the vantage point of dentists and other DHP during common dental procedures, 2) compare the effectiveness of commonly used aerosol mitigation interventions including HVE, and 3) establish the duration over which aerosols persist in the dental clinic environment following an aerosol-generating procedure.

## 2. Materials and Methods

We collected data on aerosols generated during procedures in pediatric dentistry, general dentistry, orthodontic, periodontic, and endodontic clinics in St. Louis, Missouri from July to October 2020. Aerosol sensors were used to collect data. Wi-Fi Hotspots were deployed to transmit data from dental clinics to the “cloud,” allowing the research team to monitor environmental aerosol concentrations in real-time.

### 2.1 Experimental System

#### 2.1.1 Dental Clinics

Each dental clinic had a different configuration. Pediatric and general dental operatories had a single room layout. Endodontic and periodontic clinics had semi-private operatories with partial wall barriers between dental chairs. The orthodontic clinic included a large multi-operator clinic space (approximate dimensions-35m x 20m x 20m). In each dental clinic setting, the patient was accompanied by a dentist and up to two additional DHP depending on the complexity of the procedure to be performed. Supplemental Figure 1 shows a typical dental clinic setting with the location of the aerosol sensors used for the study.

Dental procedures evaluated include: high speed drilling during debonding of orthodontic brackets; enamel and dentin cutting during cavity and crown preparation; slow speed drilling for finishing cavity preparation, polishing, and trimming during crown preparation; removal of dentin and soft tissues during endodontics; and ultrasonic scaling during teeth cleaning. Dental suction was used in all configurations with 8.2 mm tip with flow rate 74 standard cubic feet per minute at 7.0 Hg (2095.44 LPM; Henry Schein 1400 RAMVAC standard model).

#### 2.1.2 Instrumentation

A MINIMA wearable particulate matter (PM) sensor (Applied Particle Technology, St. Louis, Missouri, USA) worn on the chest pocket of the dentist’s coat was used to measure the PM concentration exposure of dentists. [16,17]. Dental procedure start and end times were recorded by dental clinic staff. The PM sensors recorded every 15 seconds; data was transmitted wirelessly in real-time to a cloud-based dashboard for visualization and temporal analysis. The number concentration of aerosols generated during each dental procedure was also measured using an optical aerosol spectrometer (Model 11C, GRIMM Aerosol Technik, Ainring, Germany) that allows particle detection in the size range from 220 nm to 32 µm [18]. The inlet of the optical aerosol spectrometer was positioned 20 cm in front of and 10 cm above the patient’s mouth without interruption of the procedures performed by the dentist.

#### 2.1.3 Test Plan

The test plan (Table 1) consisted of the four objectives for measuring the aerosol emission during the following dental procedures: high-speed drilling; slow-speed drilling; prophylaxis; and ultrasonic scaling in different dental clinic settings.

**Table 1:** List of experiments of aerosol measurements for the study

| TEST # | Objective: | Description | Dentistry Type | Result |
| --- | --- | --- | --- | --- |
| I | The effect of type of dental procedure on aerosol emission using tip HVE mitigation | 1.High-speed drill used for 10 minutes.<br>2.Slow-speed drill used for 2 minutes.<br>3.Prophy used for 5 minutes.<br>4.Ultrasonic scaler used for 15 minutes. | (A) Orthodontic<br>(B) Pediatric<br>(C) Endodontic<br>(D) Periodontic | Figure 1 |
| II | Effect of aerosol mitigation strategy on aerosol emission during operation on patient | 1.Use of high-speed drill with minimal ventilation in dental space<br>2. Use of high-speed drill with Conical HVE<br>3.Use of high-speed drill with tip HVE<br>4. Use of ultrasonic scaler with tip HVE<br>5. Intermittent use of Ultrasonic scaler with isovac<br>6. Continuous use of Ultrasonic scaler with isovac | (A)Orthodontic<br>(D)Periodontic | Figure 2 |
| III | Temporal variation of aerosol emission during operation on patient | 1.High speed drill | (A) Orthodontic<br>(E) General | Figure 3 |
| IV | Effect of operation location in oral cavity on aerosol emission | 1.High-speed drill with tip HVE on anterior teeth<br>2. High-speed drill with tip HVE on posterior teeth<br>3. Ultrasonic scaler with tip HVE on anterior teeth<br>4. Ultrasonic scaler with tip HVE on posterior teeth | (A) Orthodontic<br>(B) Pediatric<br>(D) Periodontic | Supplemental Figure 3 |

### 2.2 Data Analysis

The number concentration obtained from the optical aerosol spectrometer was used to generate the size number distribution of aerosols emitted during each dental procedure [19]. The number concentration used for the size distribution function calculation was the difference between the number concentration measured during the procedure and the number concentration measured before the start of procedure (background data).

The size distribution function of the aerosols 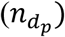 was calculated using the following equation:

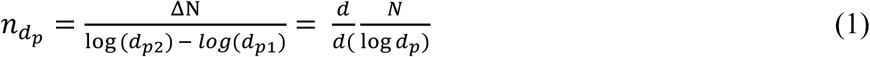

ΔN is the number concentration of the particles measured by optical aerosol spectrometer for a particular size range in the range d_p1_to d_p2_. The size distribution of aerosols is important since it estimates the number of particles in a desired size range. The total number concentration of particles in a specific size range is obtained by integrating the size distribution function over infinitesimal small particle size with the upper and lower limit as maximum and minimum particle size.

## 3. Results

### 3.1 The effect of type of dental procedure on aerosol emission using tip HVE mitigation

Figure 1 represents the size distribution of the aerosols generated during the use of high-speed drilling in orthodontics, pediatric dentistry, and endodontics, and use of an ultrasonic scaler in periodontics during routine patient care. All procedures utilized standard tip HVE (Supplementary Figure 2a) to mitigate aerosol spread in the environment. The data represents the average number concentration of aerosols emitted during the entire duration of each procedure using the dental handpiece on a patient as a function of log of the size of the aerosols. Data from use of slow-speed drill and prophylaxis are not presented as no aerosols were observed. Use of high-speed drill in orthodontics demonstrated emission of the highest concentration of particles at two modes (0.28 µm and 0.35 µm), whereas the remaining dental procedures only showed a single mode at 0.28 µm. A large number of aerosols were generated in orthodontics for the particle size less than 1µm as compared to pediatrics, endodontics during the use of high-speed drill and periodontics during the use of ultrasonic scalar.

**Figure 1.**
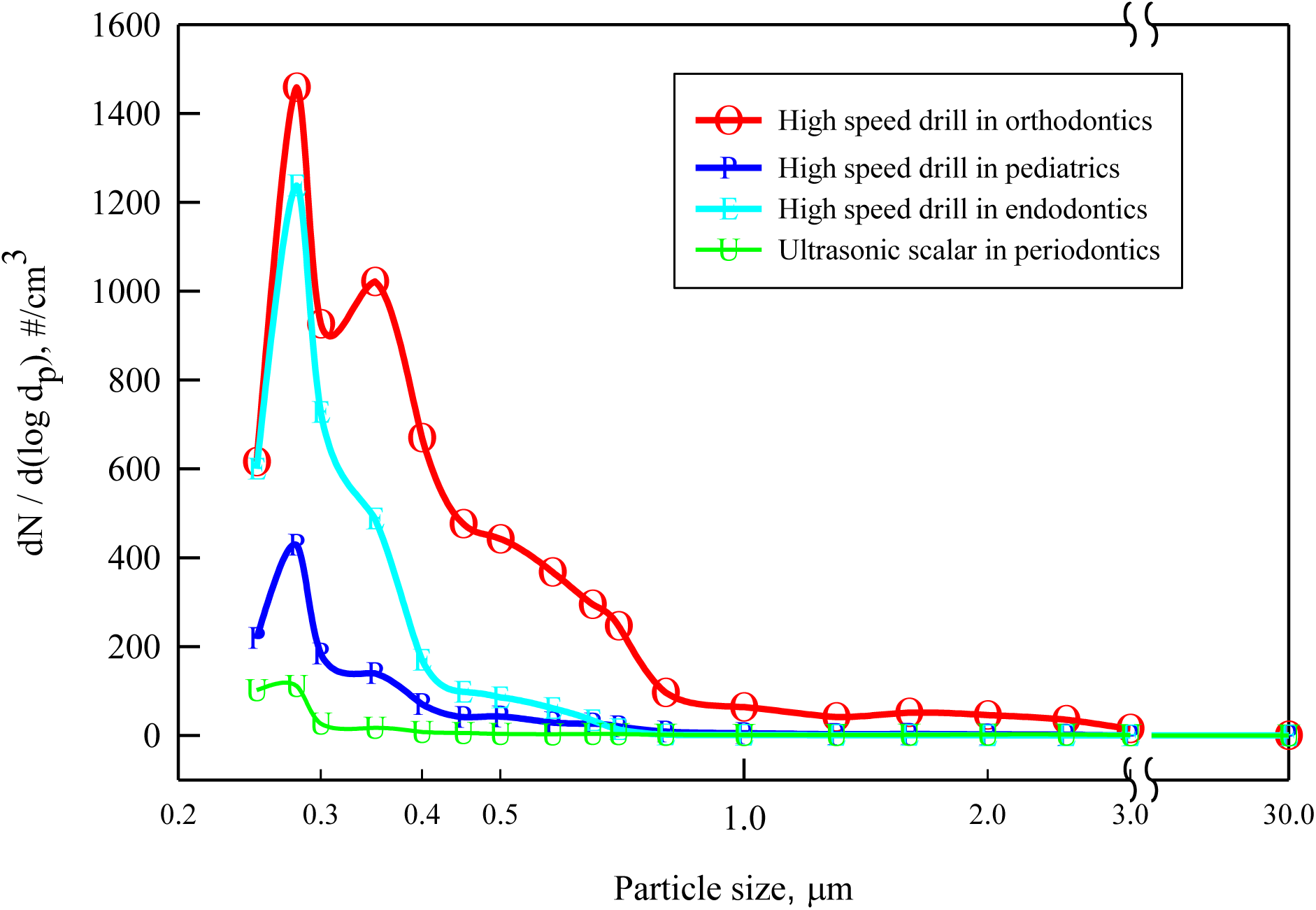
Size distribution of aerosols generated during different dental procedures using tip HVE mitigation: The concentration and size distribution of aerosols generated by slow-speed drilling in pediatrics and the use of prophylaxis in pediatrics were also measured but were close to background levels, and are therefore not shown for reasons of clarity.

Aerosol emissions from high-speed drilling in orthodontics were higher than from high-speed drilling in endodontics and pediatrics. The average concentration of particles in the size-range 250nm to 1µm that were emitted was as high as 340 particles per cm^3^ of air during 10 minutes of high-speed drilling in orthodontics (the number concentration the particle is obtained by multiplying the size distribution function with the desired size range as per the equation1), 170 particles per cm^3^ of air during 10 minutes of high-speed drilling in endodontics, 60 particles per cm^3^ of air during 10 minutes of high-speed drilling for in pediatrics and 15 particles per cm^3^ of air ultrasonic scaling for 15 minutes in periodontics.

### 3.2 Effect of aerosol mitigation strategy on aerosol emission during operation on patient

Results of aerosol emission comparing different mitigation strategies employed by dentists are presented in Figure 2. The concentration of aerosol was highest when using a high-speed drill in a dental operatory space with little ventilation, Conical HVE (Supplementary Figure 2b) was found to have higher efficiency for removing aerosol plumes compared to standard tip HVE.

**Figure 2.**
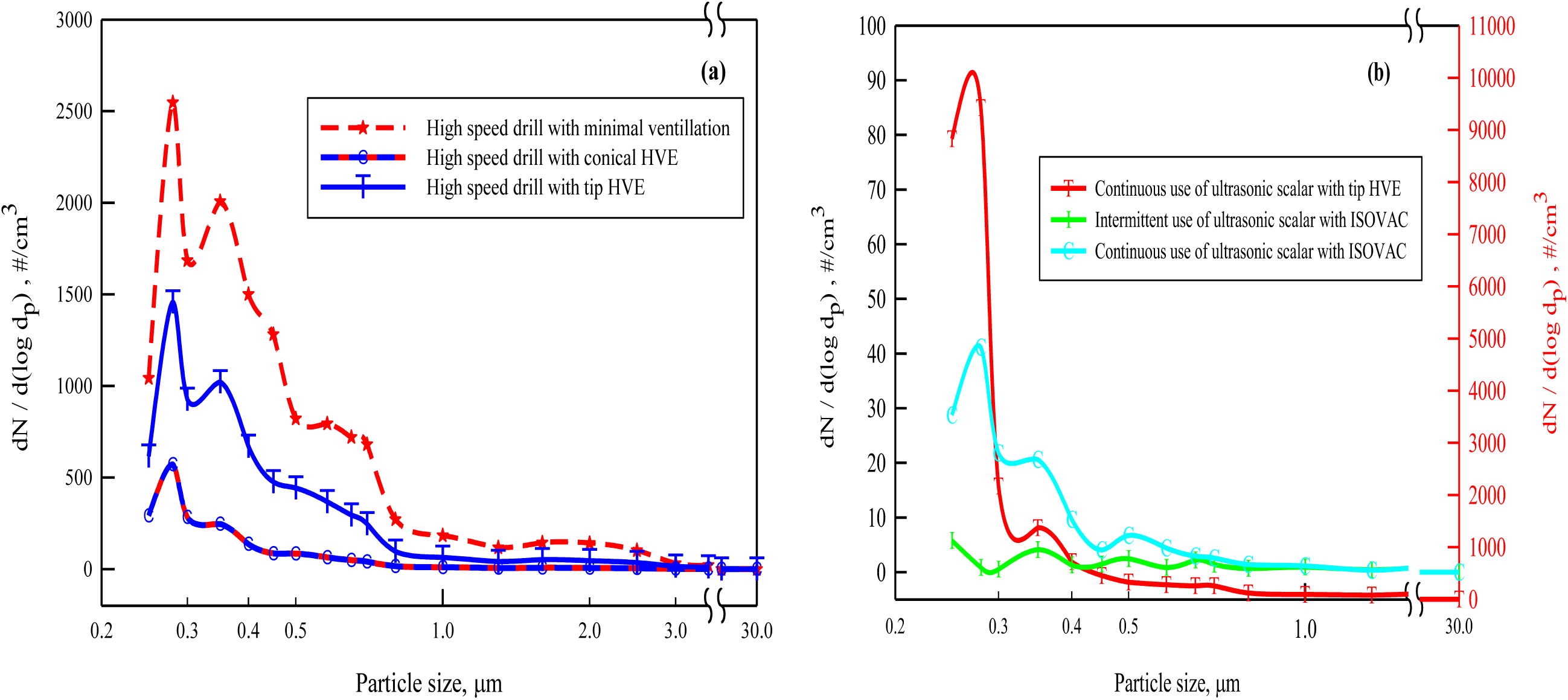
Size distribution of the aerosol emissions measured using different aerosol mitigation strategies.

The ISOVAC® (Supplementary Figure 2c) dental isolation adapter used with HVE was also evaluated during ultrasonic scaler use. Compared to standard tip HVE, ISOVAC® reduced aerosol emissions by a factor greater than 1000 during intermittent ultrasonic scaler use, and when continuous scaling was performed with ISOVAC® HVE aerosol emissions were very low but higher than intermittent ultrasonic use during the procedure.

### 3.3 Temporal variation of aerosol emission during operation on patient

The real-time variation in PM_2.5_ concentration was measured using portable low-cost PM sensors. PM_2.5_ refers to the concentration of particles with a diameter <2.5 µm, considered to be of greatest health concern [19] and most likely to be inhaled into the lower respiratory tract.

Figure 3b depicts PM_2.5_ concentration during a procedure performed in a closed single patient room. During this observation, the dentist was exposed to PM_2.5_ concentrations as high as 350 µg/m^3^ when HVE was partially used during the procedure. In contrast, Figure 3a demonstrates lower PM_2.5_ concentrations (peak ∼50 µg/m^3^) during high-speed drilling in a large open multi-operator space.

**Figure 3.**
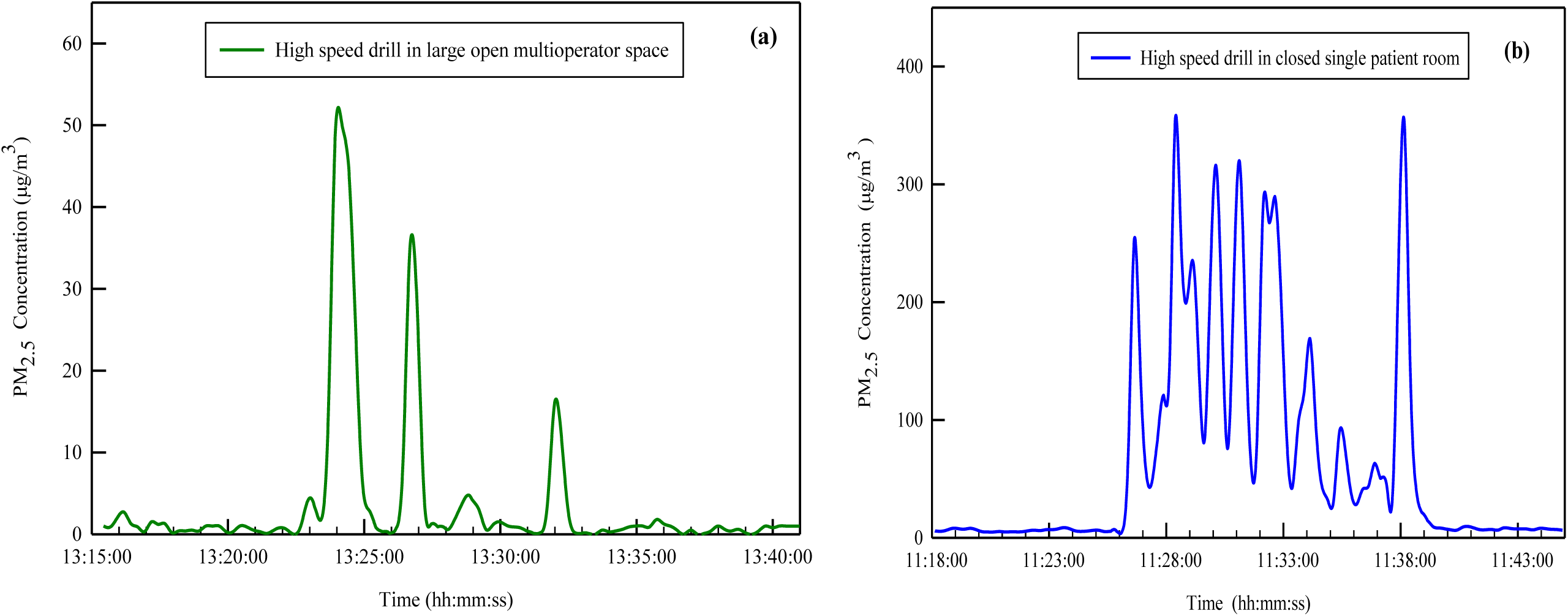
Temporal variation of PM_2.5_ concentration aerosols before, during and after dental procedures. (Note the different y-axis in two images)

## Discussion

To our knowledge, this is the first study to evaluate continuous, real-time monitoring of PM concentrations during real-world dental procedures. By using portable wearable aerosol sensors on DHPs and an optical aerosol spectrometer placed within close proximity to the patient, this study provides the most accurate representation of DHP aerosol exposure to date.

The largest aerosol concentrations were generated by ultrasonic scaling and high-speed drilling of anterior teeth (Supplementary Figure 3). This was evident during dental procedure on upper front teeth, since a huge amount of water is splashed back into the atmosphere surrounding patient and DHP with large space available for HVE to reduce the aerosols resulting in reduced efficiency of aerosol capture. In comparison, the other locations in the mouth more likely limit the spread of aerosols with use of HVE.HVE significantly reduced the aerosol concentration during all types of dental procedures sampled. Conical HVE was more effective in reducing aerosol concentrations than standard tip HVE, but ISOVAC® HVE was the most effective form of aerosol mitigation. Procedures performed in a closed single patient room demonstrated slightly higher concentrations of aerosols than those performed in larger, open and ventilated clinic spaces. However, in general, we also observed that dental aerosols, when present, appeared transient – regardless of dental clinic configuration. This indicates that fallow time can be reduced to 5 minutes – which likely occurs during routine patient care.

Further investigation of dental aerosols remains necessary. Even with aggressive aerosol mitigation interventions, dentists and other DHPs remain exposed to a small concentration of aerosols. Additionally, the size of the emitted aerosols were less than 1µm in size, well within the region where lower respiratory tract deposition could occur. Such aerosols could pose an infectious disease threat to DHPs should they harbor respiratory pathogens. Preliminary data swabbing splatter from the environment in dental settings identified SARS-CoV-2 [20]. However, this study measured dental aerosols to a limited extent, aerosols less than 10 µm can remain suspended in air for several hours before depositing on surfaces [21] and thus pathogens may still be present in the air. This study also did not use microbiologic culture to determine viability of pathogens detected. Concurrent measurement of pathogen concentrations in aerosol emissions needs to be conducted to provide a more accurate evaluation of infection risk during dental procedures.

A bimodal distribution of particle size was seen in some situations (e.g., orthodontic high-speed drilling) suggesting the possibility of two different sources of particles. The larger particle sizes measured during procedures primarily consisted of water droplets, and these were also visually observed. Further chemical characterization is necessary to differentiate between aerosols emanating from the patient and those from dental equipment. A recently published study also measured the size distribution of aerosols during different dental procedures and found that majority of particles were in the range 52 nm-72 nm at a sampling height of 60 cm [22]. However, larger droplets and particles were not found. Emissions varied from patient to patient. This highlights the importance of real-time monitoring of aerosol emissions. Slow-speed drilling and prophylaxis did not emit significant aerosols, and thus HVE may not be required during these procedures. However, this observation was made under conditions where the patient was not coughing, which is known to generate aerosols [23].

Conical HVE is likely more efficient in reducing emissions from high-speed drilling than standard tip HVE because of the relatively large surface area available for conical HVE to evacuate aerosols from the dental environment. Our data demonstrated that the addition of ventilation further prevented accumulation of aerosols in the dental environment. This was largely due to the air dilution effect and transportation of aerosols away from the immediate dental setting. Use of ISOVAC® HVE during ultrasonic scaling provided better aerosol mitigation than standard tip HVE. Aerosol mitigation of ultrasonic scaling procedures was best when ISOVAC® HVE and an intermittent scaling approach were used together. This helped by providing enough time for aerosols generated to be evacuated. DHPs who anticipate performing ultrasonic scaling for a prolonged period of time may wish to turn off the scaler intermittently during routine care to allow aerosols to dissipate.

## 4. Conclusions

Using real-world data obtained from wearable aerosol sensors and an optical aerosol spectrometer to approximate DHP exposure to aerosols during routine care, we observed that intraoral HVE was the most efficient aerosol mitigation strategy followed by conical suction and standard tip suction. Despite these interventions, dentists and other DHP remain exposed to aerosols which may place them at higher risk for respiratory infection. However, the duration of exposure can be reduced with intermittent utilization of aerosol generating devices. Furthermore, when observed, aerosols appeared to be transient and localized near the patient’s chair. These findings suggest that policies requiring dental chairs to remain fallow between patients may not be necessary.

## Data Availability

The preprint data is available to readers on request from corresponding author.

## Acknowledgements

Research in this publication was supported by the National Institute of Dental and Craniofacial Research (NIDCR) within the National Institutes of Health under award numbers U19-DE-28717 and U01-DE-28727 sub awards X01-DE-030402, X01-DE-030403, and X01-DE-031119; award K23-DE-029514; and the Washington University Institute of Clinical and Translational Sciences which is, in part, supported by the NIH/National Center for Advancing Translational Sciences (NCATS), CTSA grant UL1-TR-002345. In addition, this study was also supported in part by The Foundation for Barnes-Jewish Hospital and their generous donors. The content is solely the responsibility of the authors and does not necessarily represent the official views of the National Institutes of Health. The authors would like to also acknowledge the NIDCR practice based research network directors, Drs. Gregg Gilbert and Mary-Ann McBurnie, advisory board members, and Dr. Dena Fischer for their guidance and review of our preliminary data.

## References

1. SK Harrel, J. Molinari, Aerosols and splatter in dentistry: A brief review of the literature and infection control implications, JADA, 135(4) (2004), pp.429–437. https://doi.org/10.14219/jada.archive.2004.0207

2. S Dutil, A Mériaux, M-C de Latrémoille, L Lazure, J Barbeau, C. Duchaine, Measurement of airborne bacteria and endotoxin generated during dental cleaning. JOccup and Environ Hyg. 6(2) (2008), pp.121–130. https://doi.org/10.1080/15459620802633957

3. PS Kumar, K. Subramanian. Demystifying the mist: Sources of microbial bioload in dental aerosols. J periodontology 91(9)(2020), pp.1113–1122. https://doi.org/10.1002/JPER.20-0395

4. AM Bennett, MR Fulford, JT Walker,DJ Bradshaw, MV Martin, PD Marsh. Microbial aerosols in general dental practice. BrDental J. 189(12) (2000), pp.664–667. https://doi.org/10.1038/sj.bdj.4800859

5. SK Harrel, JB Barnes, F Rivera-Hidalgo. Aerosol and splatter contamination from the operative site during ultrasonic scaling. JADA, 129(9)(1998), pp.1241–1249. https://doi.org/10.14219/jada.archive.1998.0421

6. MF Timmerman, L Menso, J Steinfort,AJ van Winkelhoff, GA van der Weijden. Atmospheric contamination during ultrasonic scaling. Jclin periodontology. ;31(6)(2004), pp.458–462. https://doi.org/10.1111/j.1600-051X.2004.00511.x

7. V Checchi, P Bellini, D Bencivenni, U. Consolo. Covid-19 dentistry-related aspects: A literature overview. International Dental Journal Covid-19 interactive map. 2020 https://coronavirus.jhu.edu/map.html. [accessed 2020].

8. JB Epstein, K Chow, R Mathias R. Dental procedure aerosols and covid-19. Lancet Infect Dis.21 (4) (2020).e73, https://doi.org/10.1016/S1473-3099(20)30636-8

9. L Meng, F Hua, Z Bian. Coronavirus disease 2019 (covid-19): Emerging and future challenges for dental and oral medicine. JDR.;99(5)(2020),pp.481–487. https://doi.org/10.1177/0022034520914246

10. A Gioda, G Hanke, A Elias-Boneta, B Jiménez-Velez. A pilot study to determine mercury exposure through vapor and bound to pm10 in a dental school environment. ToxicolInduHealth.; 23(2)(2007), pp.103–113.

11. CG Helmis, J Tzoutzas, HA Flocas, CH Halios, OI Stathopoulou, VD Assimakopoulos, V Panis, M Apostolatou, G Sgouros, E. Adam. Indoor air quality in a dentistry clinic. Scie Total Environ.; 377(2)(2007), pp.349–365. https://doi.org/10.1016/j.scitotenv.2007.01.100

12. J Balanta-Melo, A Gutiérrez, G Sinisterra, MdM Díaz-Posso, D Gallego, J Villavicencio,A. Contreras. Rubber Dam Isolation and High-Volume Suction Reduce Ultrafine Dental Aerosol Particles: An Experiment in a Simulated Patient. Appl Sci. 10(18)(2020), pp.6345. https://doi.org/10.3390/app10186345

13. T Eliades, D Koletsi. Minimizing the aerosol-generating procedures in orthodontics in the era of a pandemic: Current evidence on the reduction of hazardous effects for the treatment team and patients. Am J Orthod Dentofacial Orthop.; 158(3)(2020) pp.330-342. https://doi.org/10.1016/j.ajodo.2020.06.002

14. C Hallier, DW Williams, AJC Potts, MAO Lewis. A pilot study of bioaerosol reduction using an air cleaning system during dental procedures. Bri dental j.; 209(8)(2010) e14 https://doi.org/10.1038/sj.bdj.2010.975

15. Richard Holliday, James R. Allison, Charlotte C. Currie, David C. Edwards, Charlotte Bowes, Kimberley Pickering, Sarah Reay, Justin Durham, Joanna Lumb, Nadia Rostami, Jamie Coulter, Christopher Nile, Nicholas Jakubovics, Evaluating contaminated dental aerosol and splatter in an open plan clinic environment: Implications for the COVID-19 pandemic, J Dentistry,105(2021),103565. https://doi.org/10.1016/j.jdent.2020.103565

16. J. Li, SK Mattewal, S Patel, P Biswas. Evaluation of nine low-cost-sensor-based particulate matter monitors.AAQR, 20(2)(2020), pp.254270. https://doi.org/10.4209/aaqr.2018.12.0485

17. Applied particle technology. https://appliedparticletechnology.com/. [accessed 2020 September 10].

18. S. Sousan, K Koehler, L Hallett, TM Peters. Evaluation of the alphasense optical particle counter (opc-n2) and the grimm portable aerosol spectrometer (pas-1.108). Aerosol Sci Tech.; 50(12)(2016) pp.1352-1365. https://doi.org/10.1080/02786826.2016.1232859

19. W.C. Hinds. Aerosol technology: Properties, behavior, and measurement of airborne particles. John Wiley & Sons; Hoboken, NJ, USA. 1999

20. AP Meethil, S Saraswat, PP Chaudhary,SM Dabdoub,PS Kumar. Sources of SARS-CoV-2 and Other Microorganisms in Dental Aerosols. J Dent Res. 100(8) (2021)817–823. https://doi.org/10.1177/00220345211015948

21. Dhawan, S. and P. Biswas, Aerosol Dynamics Model for Estimating the Risk from Short-Range Airborne Transmission and Inhalation of Expiratory Droplets of SARS-CoV-2. Environmental Science & Technology, 2021. 55(13): p. 8987–8999. https://doi.org/10.1021/acs.est.1c00235

22. B. Polednik. Exposure of staff to aerosols and bioaerosols in a dental office. Building and Environment. 187(2021).107388. https://doi.org/10.1016/j.buildenv.2020.107388

23. Klompas, M., M. Baker, and C. Rhee, What Is an Aerosol-Generating Procedure? JAMA Surgery, 2021. 156(2).pp. 113–114 https://doi.org/10.1001/jamasurg.2020.6643

