## Supplement for "A Comparison of Aerosol Mitigation Strategies and Aerosol Persistence in Dental Environments"

### (SUPPLEMENTARY APPENDIX)

APPENDIX

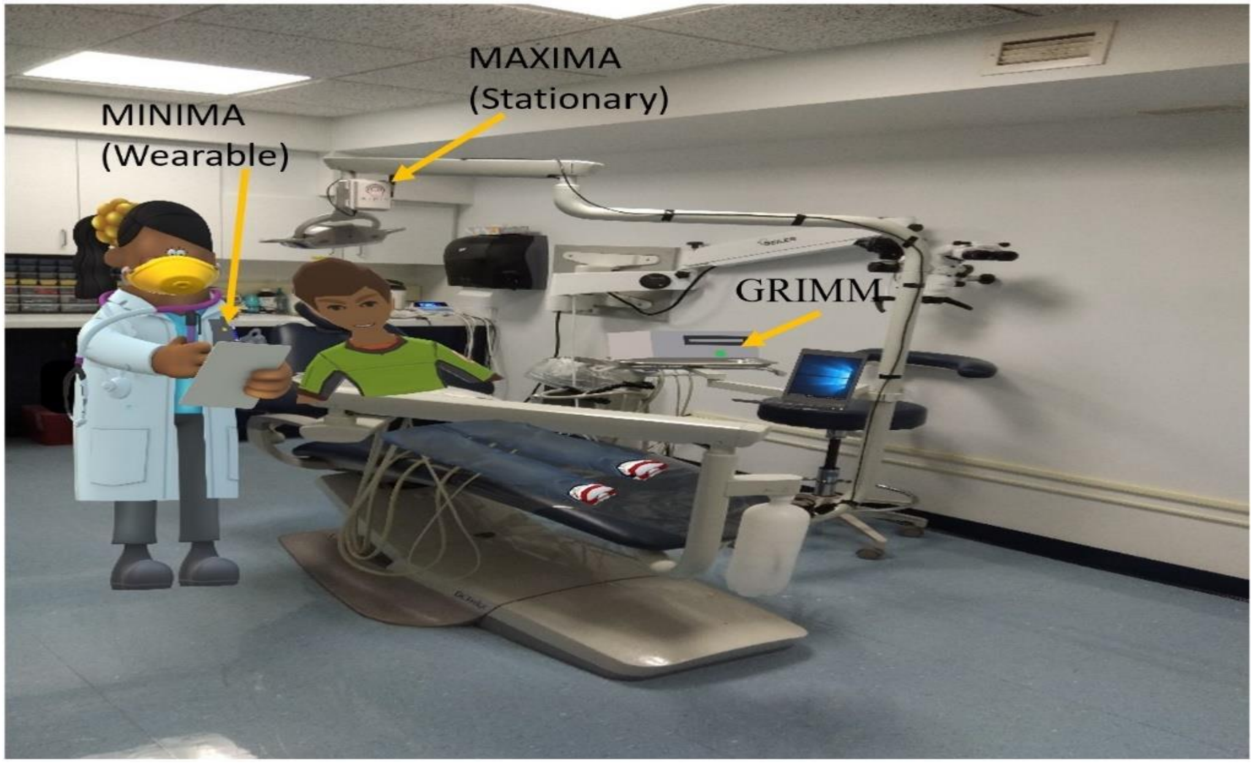


*Figure 1. Position of dentist, patient, MAXIMA, MINIMA and GRIMM aerosol detection instruments in the dental setting*

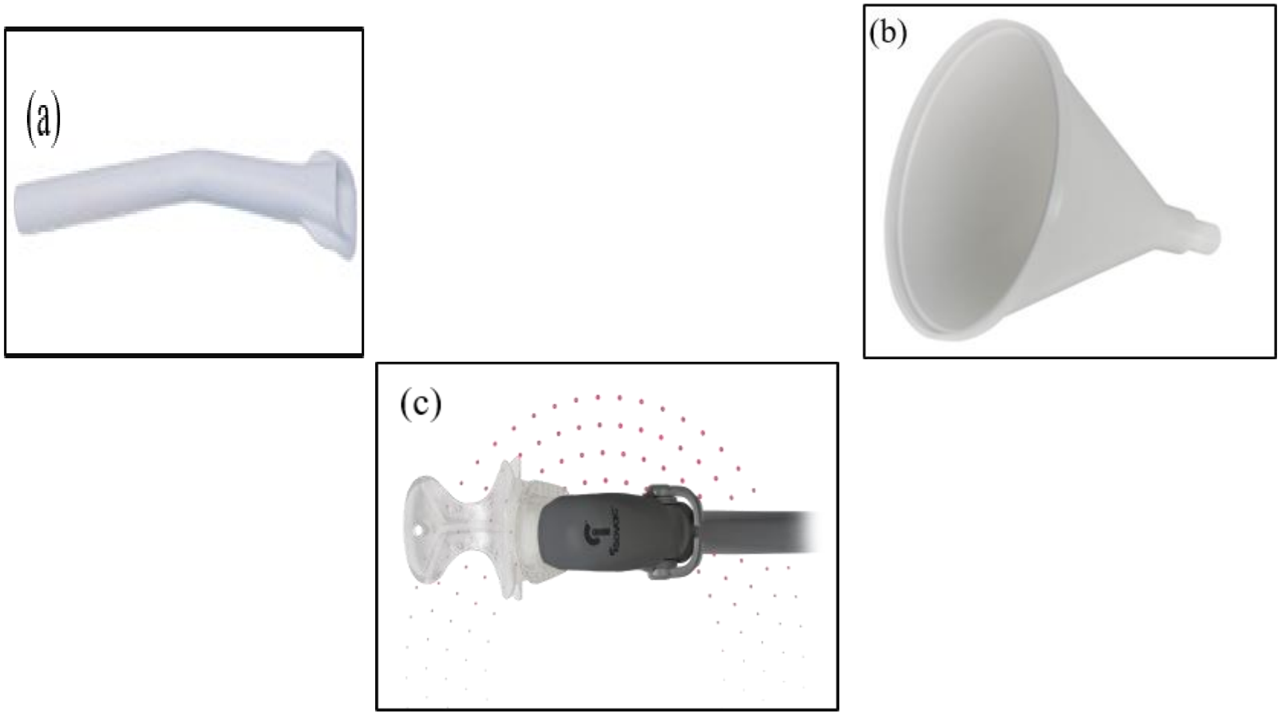


*Figure 2 (a) Tip HVE, (b) Conical HVE, (c) Isovac® dental isolation adaptor for use with HVE*

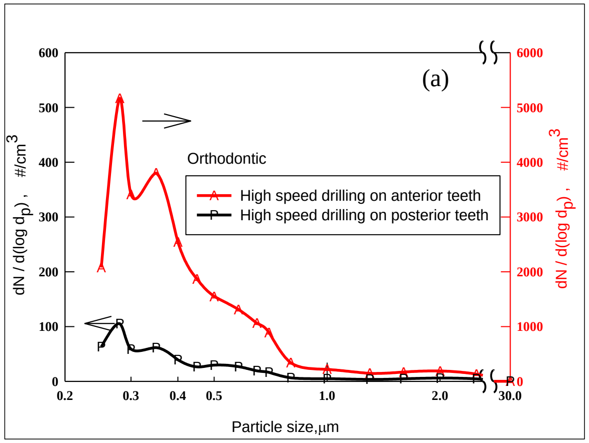
(a)
Orthodontic
High speed drilling on anterior teeth
High speed drilling on posterior teeth
dN / d(log $d_p$) , #/cm$^3$
Particle size,µm
0
100
200
300
400
500
600
1000
2000
3000
4000
5000
6000
0.2
0.3
0.4
0.5
1.0
2.0
30.0

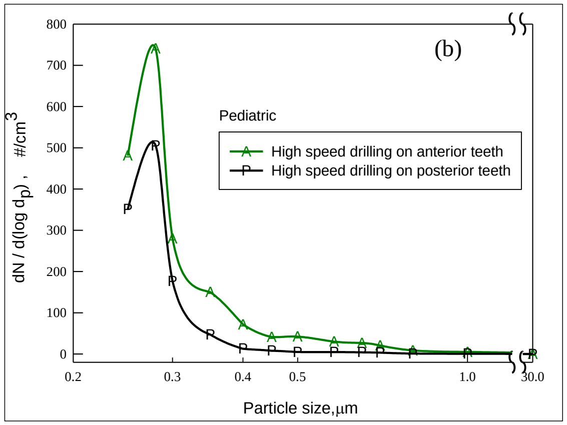
(b)
Pediatric
High speed drilling on anterior teeth
High speed drilling on posterior teeth
dN / d(log $d_p$) , #/cm$^3$
Particle size,µm
0
100
200
300
400
500
600
700
800
0.2
0.3
0.4
0.5
1.0
30.0

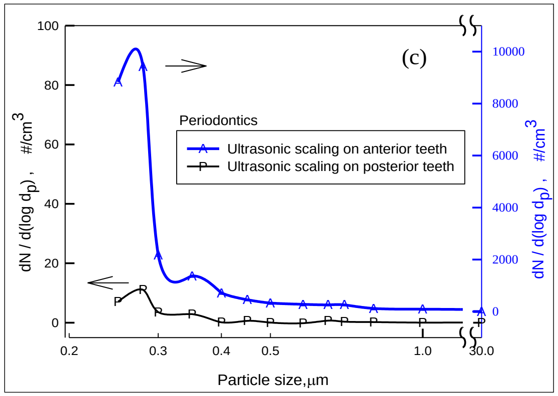
(c)
Periodontics
Ultrasonic scaling on anterior teeth
Ultrasonic scaling on posterior teeth
dN / d(log $d_p$) , #/cm$^3$
Particle size,µm
0
20
40
60
80
100
2000
4000
6000
8000
10000
0.2
0.3
0.4
0.5
1.0
30.0

*Figure 3 Size distribution of the aerosol emissions during operation on anterior and posterior teeth in (a) Orthodontics during use of high speed drill, (b) pediatrics during use of high speed drill, (c) periodontics during use of ultrasonic scalar*
